# EXPLORING WOMEN’S PERSPECTIVES ON ABORTION LEGALIZATION IN NAMIBIA: INSIGHTS FROM RESIDENTS OF /GERERES, KEETMANSHOOP

**DOI:** 10.1101/2023.07.20.23292972

**Authors:** Rauna Namukwambi, Lovis Sheehama, Justice EK. Sheehama, Hilma N. Nakambale

## Abstract

**Introduction:** Abortion is a globally controversial topic. Despite Namibia gaining independence in 1990, the country still upholds the apartheid inherited 1975 Abortion and Sterilization Act of South Africa, which places restrictions on abortion. There have been calls from many parliamentarians to legalize abortion on demand due to the alarming rate of backyard abortions that were putting Namibian women’s lives at risk. However, there is opposition from Pro-life advocates. This study assessed the perceptions of women in /Gereres, Keetmanshoop regarding the legalization of abortion in Namibia.

**Methods:** This study utilized a quantitative, descriptive cross-sectional design. A non-probability systematic sampling technique was used to select participants. A self-administered questionnaire was used to collect data. Participants’ perspectives were captured using structured three-point and five-point Likert scale checklists. Microsoft Excel was used to analyze data. Data were presented using frequency tables.

**Results:** The study revealed a variation of views from participants, mostly opposing the legalization of abortion in Namibia. Most women acknowledged that illegal abortion poses risks to women; however, they would not advocate for the legalization of abortion. Notably, several participants appeared to justify abortion in certain circumstances such as in cases of rape or incest, or if continuation of the pregnancy poses a health risk to the woman. Cultural and religious beliefs appeared to be major factors influencing women’s opposing views on abortion.

**Conclusion:** The study highlights the complex and diverse perspectives of women in /Gereres, Keetmanshoop regarding the legalization of abortion in Namibia. While recognizing the risks associated with illegal abortion, most participants opposed its legalization, which could mostly be attributed to cultural and religious beliefs. However, some participants acknowledged specific circumstances where abortion could be justified. This study provides women’s insights on the legalization of abortion in Namibia, useful for lawmakers, policymakers, and stakeholders in the field of reproductive health.

## 1. Background

Abortion is a highly contentious topic that permeates global cultural, social, moral, religious, and legal facets of life (1). Over 50 years ago, the World Health Assembly recognized abortion as a serious public concern (2). Addressing abortion as a national and global public health imperative should therefore be at the forefront of law and public policy deliberations as unsafe abortions continue to end the lives of many young women globally (3).

In Namibia, abortion is only permissible under special circumstances, as outlined in the Abortion and Sterilization Act of 1975. These circumstances include when the pregnancy is a danger to the woman’s physical and mental health, pregnancy due to rape and incest, and other cases (4). Although South Africa, from which Namibia inherited the abortion law post-apartheid, changed its abortion law in 1996, Namibia continues to criminalize abortion even after its independence (5). Attempts to revise the Namibian Abortion and Sterilization Act of 1975 have been met with strong opposition from religious leaders and women’s groups (5–10). The bill to oppose the Abortion and Sterilization Act of 1975 was first issued in 1996 by Namibia’s first Minister of Health and Social Services for public consultation. The bill aimed to replace the apartheid-inherited South African Abortion and Sterilization Act of 1975 and to extend the legality of abortion where a procedure can be done on demand within a specific time frame. However, the bill was ultimately opposed in 1999 due to severe public scrutiny (11).

Recently, an online petition which was launched to oppose the legalization of abortion in Namibia gathered more than 15,000 signatures (5). Several petitions on both sides of the abortion debate have since been tabled in parliament; however, the motion is yet to be debated and voted on (6). Therefore, to date, abortion in any other circumstances outside the current Abortion and Sterilization Act of 1975 special provisions remains a criminal offence for women and persons who perform it. The punishment for illegal abortion is a fine of up to N$5,000 (∼USD 300) or an imprisonment term of up to five years, or both (12).

Although abortion is massively restricted in Namibia, this does not prevent women and girls from terminating unwanted pregnancies. The situation has forced many women to resort to unsafe abortions, contributing to maternal death (12–15). Statistics on women who opt for abortion in Namibia are hard to obtain as many cases go undocumented (16). The local media reported that in 2016, the country recorded 7,335 suspected cases of illegal abortions in state healthcare facilities (17).The Namibian police’s statistics between 2013 to 2019 have only confirmed 2% of the 138 authorized abortion cases reported (16).

There is currently limited knowledge of the perceptions of women regarding the legalization of abortion on demand and the factors contributing to such perceptions. Therefore, this study assessed the perceptions of women residing in /Gereres, Keetmanshoop, on legalizing abortion in Namibia, as women experience the effects of the present laws on abortion.

## 2. MATERIALS AND METHODS

### 2.1 Research design

The study employed a quantitative cross-sectional survey method. The survey explored the perceptions of women on how the legalization of abortion would affect the demand for abortion services.

### 2.2 Study setting and population

This study was conducted at /Gereres location in the Keetmanshoop district. Keetmanshoop is a town in the southern part of Namibia, in the //Karas region, with a population of approximately 26,000 according to the latest population census data (18). The study population consisted of women of childbearing age residing in /Gereres location, Keetmanshoop, between the ages of 18 to 49 years. Male participants, women below the age of 18, and individuals who could not consent to participate were excluded from the study.

### 2.3 Sample and sampling method

Given the relatively small population size of our study setting, the survey targeted all women ages 18-49 years. A combination of purposive and systematic sampling was used to determine the sample size. The first participant was randomly selected as a starting point; after that, the researcher approached every second house to recruit participants. There was no specific reason for selecting this method; however, the researchers felt it was in the interest of the study findings to gather data. The sample size for this study was forty women.

### 2.4 Research Instruments

The study used a structured self-administered questionnaire consisting of close-ended questions. The questionnaire was constructed in English and translated into Afrikaans, Rukwangali, and Oshiwambo as these were the languages spoken by most of the participants in that area. The questionnaire consisted of two sections. Section A included socio-demographic data of the participants (age, gender, religion, marital status, language, and educational qualifications); Section B was on perceptions regarding the legalization of abortion, and participants were asked to state their opinions using a 3-point Likert scale requiring a “yes”, “no” or “not sure” and 5-point Likert scale ranging from 1=strongly disagree to 5= strongly agree. The questionnaire was constructed based on the literature on abortion, specifically from the Namibian and Sub-Saharan Africa context.

### 2.5 Validity and Reliability

The questionnaire was examined for validity by experts in the field of reproductive health, in the Department of Reproductive Health at the Ministry of Health and Social Services, to assess the extent to which the questions were going to fulfill the aims of the study. To enhance the reliability of the questionnaire, a pilot study was conducted to pre-test the study instrument to ensure consistent results. The questionnaire was piloted twice on seven random participants in Kopeislaagte location, Keetmanshoop, to confirm reliability (the findings from the pilot study are not part of the findings in this study).

### 2.6 Procedures for data collection

The primary researcher approached potential participants, explained the purpose of the study, and obtained consent from participants to proceed with the study. Participants were recruited between August and September 2022. Data collection took eight to fifteen minutes for each participant to complete the questionnaire. The data collection process took three weeks. The researcher wore a mask and used hand sanitizer for herself and the participants to ensure COVID-19 regulations were adhered to. A social distance of about 1.5 meters was also put in place.

### 2.7 Data analysis

After data collection, responses to the questionnaire were entered into a Microsoft Excel 2019 sheet, and a statistician analyzed and summarized the data. Descriptive statistics, including frequency tables, were used to present the findings.

### 2.8 RESEARCH ETHICS and Informed Consent

Ethical approval was obtained from the School of Nursing and Public Health’s Ethics Committee, before commencing with the study (**Annexure A**). The municipality of Keetmanshoop also issued a letter of approval to conduct the study. The research was conducted on human beings; thus, the researcher was guided by the Belmont Ethical Principles and Guidelines for the Protection of Human Subjects of Research (19).

Full informed consent was issued to the participants. The researcher explained the purpose of the study and research objectives to the participants, informed them about their rights to refuse to participate, and ensured that participants understood the information to decide whether to consent. Participants consented to participate by signing the informed consent form (**Annexure B**) before completing the questionnaire.

## 3. RESULTS

The demographic characteristics of the study participants are shown in **Table 1**. A total of 40 participants were included in the study. Majority of the participants (55.0%) were between the ages of 18-27 years. Most participants were single (65.0%), with 42.5% having completed grades 8-10 as their highest level of education. An equal number of participants (37.5%) spoke Oshiwambo or Khoekhoegowab as a home language. All participants in this study were of the Christian religion.

**Table 1.** Participant Demographic characteristics (N=40)

| Demographic characteristics | n (%) |
| --- | --- |
| **Age in years** |  |
| 18-27 | 22(55.0%) |
| 28-37 | 11(27.5%) |
| 38-47 | 6(15.0%) |
| ≥ 48 | 1(2.50%) |
| **Marital Status** |  |
| Single | 26(65.0%) |
| Living with a partner | 9(22.5%) |
| Married | 4(10.0%) |
| Other | 1(2.5%) |
| **Education level** |  |
| Grades 1 to 7 | 4 (10.0%) |
| Grades 8 to 10 | 17(42.5%) |
| Grades 11 to 12 | 15(37.5%) |
| No formal education | 4(10.0%) |
| **Home Language** |  |
| Oshiwambo | 15(37.5%) |
| Khoekhoegowab | 15(37.5%) |
| Rukwangali | 5(12.5%) |
| Afrikaans | 3(7.5%) |
| Other | 2(5.0%) |
| **Religion** |  |
| Christian | 40(100%) |

**Note:** N=40

### 3.1 Participants’ ethical, moral, and cultural perceptions regarding the legalization of abortion

Overall, most of the participants (65.0%) felt that abortion should not be legalized because it is a sin, with half of the participants in the study stating that they would not advocate for the legalization of abortion if it were against their religious beliefs. Nearly all participants (90.0%) stated that their religion prohibits women from having an abortion. Similarly, most of the participants (62.5%) stated that they are against abortions because abortions are culturally unacceptable, and that abortion constitutes murder. While most of the participants (62.5%), indicated that illegal abortions pose health risks to women, an equal proportion felt that legalizing abortion would encourage people to have more sexual partners. Notably, most participants (62.5%) indicated that they would not advocate for abortion to be legalized on demand (**Table 2**).

**Table 2.** Ethical, moral and cultural perceptions regarding the legalization of abortion.

| Perceptions | Degree of agreement<br>n (%) |  |  |
| --- | --- | --- | --- |
|  | Yes | No | Not sure |
| Abortion should not be legalized because it is a sin | 26(65.0%) | 11(27.5%) | 3(7.5%) |
| Would you advocate for the legalization of abortion if it is against your religious beliefs? | 16(40.0%) | 20(50.0%) | 4(10.0%) |
| Does your religion allow people to have an abortion? | 1(2.5%) | 36(90.0%) | 3(7.5%) |
| I am against abortion because it is not accepted in my culture | 25(62.5%) | 11(27.5%) | 4(10.0%) |
| Abortion is murder | 25(62.5%) | 13(32.5%) | 2(5.0%) |
| In your opinion, do illegal abortions pose any health risks? | 25(62.5%) | 7(17.5%) | 8(20.0%) |
| Legalizing abortion will encourage people to have more sexual partners | 25(62.5%) | 13(32.5%) | 2(5.0%) |
| Would you advocate for abortion to be legalized on demand? | 12(30.0%) | 25(62.5%) | 3(7.5%) |

**Note:** $N=40$

### 3.2 Participants legal and socio-economic perceptions regarding the legalization of abortion

Participants had mixed opinions regarding whether abortion should be legalized under different social and economic circumstances. None of the participants strongly agreed that abortion should be legalized in cases of poverty or financial problems. Moreover, participants had divided opinions on whether abortion should be allowed if a young girl was in school or too young. Some participants (37.5%) strongly disagreed that abortion should be allowed in cases where the mother does not want to keep the baby, with only 2 participants (5%) agreeing that women should be free to decide whether to have an abortion. Only 22.5% of the participants agreed that access to abortion is a woman’s reproductive right. **Table 3**

**Table 3.** Legal and socio-economic perceptions for permissible abortion.

| Perceptions | Degree of Agreement<br>n (%) |  |  |  |  |
| --- | --- | --- | --- | --- | --- |
|  | Strongly agree | Agree | Uncertain | Disagree | Strongly Disagree |
| Abortion should be allowed in cases of poverty or financial problems | 0(0.0%) | 12(30.0%) | 5(12.5%) | 18(45.0%) | 5(12.5%) |
| Abortion should be allowed if a girl is in school or is too young (below the legal age of sexual consent) | 2(5.0%) | 14(35.0%) | 5(12.5%) | 14(35.0%) | 5(12.5%) |
| Abortion should be allowed in cases where the mother does not want to keep the baby | 10(25.0%) | 7(17.5%) | 2(5.0%) | 6(15.0%) | 15(37.5%) |
| Women should be free to decide whether to have an abortion or not | 13(32.5%) | 2(5.0%) | 3(7.5%) | 4(10.0%) | 18(45.0%) |
| Access to safe abortion is a woman's reproductive right | 5(12.5%) | 9(22.5%) | 7(17.5%) | 14(35.0%) | 5(12.5%) |

**Note:** $N=40$

### 3.3 Participants’ perceptions on the legal and health conditions for permissible abortion

**Table 4** summarizes participants’ perceptions on the legal and health conditions for permissible abortion. Most of the participants (52.5%) strongly agreed that abortion should be allowed if the pregnancy was a result of rape/sexual assault or incest. Furthermore, most of the participants (65.0%) agreed that abortion should be allowed if the continuation of the pregnancy poses a health risk to the woman. Only 10% and 2.5% of participants strongly agreed that abortion should be allowed if the pregnant woman is mentally incapacitated or if the fetus has congenital anomalies, respectively.

**Table 4:** Perceptions of legal and health conditions for permissible abortion.

| Perceptions | Degree of Agreement<br>n (%) |  |  |  |  |
| --- | --- | --- | --- | --- | --- |
|  | Strongly Agree | Agree | Uncertain | Disagree | Strongly Disagree |
| Abortion should only be allowed in cases of rape/sexual assault or incest | 21(52.5%) | 12(30.0%) | 1(2.5%) | 3(7.5%) | 3(7.5%) |
| Abortion should be allowed if the continuation of the pregnancy poses a health risk to the woman | 6(15.0%) | 26(65.0%) | 4(10.0%) | 3(7.5%) | 1(2.5%) |
| Abortion should be allowed if the pregnant woman is mentally incapacitated | 4(10.0%) | 18(45.0%) | 3(7.5%) | 12(30.0%) | 3(7.5%) |
| Abortion should be allowed if the fetus has congenital anomalies | 1(2.5%) | 17(42.5%) | 8(20.0%) | 12(30.0%) | 2(5.0%) |

**Note:** $N=40$

### 3.4 Participants’ perceptions on the effects of legalizing abortion on women’s health and on the overall societal perception on abortion

When asked about the potential consequences of legalizing abortion, 35.0% and 42.5% of participants agreed that access to safe abortions may result in women/girls using abortion as a substitute for contraceptives and may result in undermining pregnancy prevention efforts, respectively. While most participants (57.5%) agreed that restrictions to safe abortions push women/girls into unsafe abortion practices, none of the participants strongly agreed that legalizing would reduce the social stigma surrounding illegal abortion and only 7.5% strongly agreed that legalizing abortion may decrease baby dumping, **Table 5**.

**Table 5:** Perceptions of the effects of legalizing abortion on women’s health and on the overall societal perception on abortion.

| Perceptions | Degree of Agreement<br>n (%) |  |  |  |  |
| --- | --- | --- | --- | --- | --- |
|  | Strongly Agree | Agree | Neutral | Disagree | Strongly Disagree |
| Access to safe abortions may result in women/girls using abortion as a substitute for contraceptives | 5(12.5%) | 14(35.0%) | 6(15.0%) | 12(30.0%) | 3(7.5%) |
| Access to a safe abortion may result in the undermining of pregnancy prevention efforts | 5(12.5%) | 17(42.5%) | 4(10.0%) | 12(30.0%) | 2(5.0%) |
| Restrictions to safe abortions push women/girls into unsafe abortion practices | 3(7.5%) | 23(57.5%) | 6(15.0%) | 6(15.0%) | 2(5.0%) |
| Legalizing abortion would reduce the social stigma surrounding illegal abortion. | 0(0.0%) | 11(27.5%) | 14(35.0%) | 9(22.5%) | 6(15.0%) |
| Legalizing abortion may decrease baby dumping | 3(7.5%) | 14(35.0%) | 8(20.0%) | 9(22.5%) | 6(15.0%) |

**Note:** *N=40*

## Discussion

The legalization of abortion remains a highly contested issue in Namibia. In this study, we explored the perceptions of women regarding the legalization of abortion in Namibia, as well as the factors influencing these perceptions. Overall, our study found that women in //Gereres, Keetmanshoop did not support the legalization of abortion. Several reasons were indicated as contributing factors to the poor support for abortion legalization. Firstly, participants found abortion to be ethically, morally, and culturally unacceptable. Most of the participants in our study indicated that abortion should not be legalized as they viewed it as a sin, and that it constituted murder. These findings are consistent with studies in Kenya and Nigeria that found that participants did not support abortion for religious and moral reasons (20,21). Similarly, a study in the UK found that most participants acknowledged that abortion was socially unacceptable and viewed as highly taboo (22). These findings were not surprising, considering that all the participants in our study were affiliated with the Christian religion. It is highly plausible that the participants’ perceptions regarding the legalization of abortion are deeply rooted in religious scriptures and doctrine, as well as perceptions and theories about when life begins. Religious doctrine and beliefs may impede public health recommendations regarding abortion (23) and may potentially propagate the prohibition of abortion in Namibia.

Despite most participants acknowledging that illegal abortions pose health risks to women, most of the participants indicated that they would not advocate for abortion to be legalized on demand. Comparative studies in Namibia found that 49% of participants stated that they would not advocate for abortion to be legalized under certain provision, despite 93% of respondents agreeing that illegal abortions pose health risks (12). These findings may indicate strong support for the current restrictive abortion laws among Namibians, regardless of the associated health implications of unsafe abortions. Therefore, alternative methods may need to be provided to reduce the health risks to women due to illegally performed abortions. Taken together, these results highlight the interplay of religion and culture in shaping perceptions regarding the legalization of abortion and are therefore important factors to be considered when assessing the acceptability of abortion in Namibian communities.

Secondly, our study found that most women believed that legalizing abortion would promote promiscuity. Similar to our findings, a study by Bisong et al (2016) reported that most of the participants in the study stated that legalizing abortion would promote promiscuity (24). In contrast, a study in Ghana that investigated the perceptions of schoolgirls regarding abortion found that most of the respondents disagreed that access to abortion services would result in women having more sexual partners (25). While these two studies did not conduct statistical analysis to assess the correlation between abortion legalization and sexual behavior, a study in the US found evidence to suggest a causal relationship between abortion legalization and increased risky sexual behavior, as well as increased incidences of sexually transmitted diseases (26). A study by Jonathan & Stratmann (2003) found that incidences of gonorrhea and syphilis were significantly and positively correlated with the legalization of abortion in the US in the 1970s (26). While these findings may not necessarily be generalizable, they provide important considerations for legalizing abortion in Namibia, given the current burden of HIV and other sexually transmitted diseases in the country (27,28). These findings highlight that the legalization of abortion may need to be followed by stringent policies to increase the accessibility and availability of contraceptives, such as condoms, to prevent the transmission of sexually transmitted diseases and other potential, unintended consequences.

We observed interesting variations in the perceptions regarding abortion when participants were provided with various case scenarios. We found that participants had divided opinions on whether abortion should be legalized in cases of poverty or financial problems, and if a girl was in school or was too young. Societal norms and economic and legal obstacles have a significant influence on women’s decisions to terminate pregnancy (29). On the permissibility of abortion in schoolgirls, several studies have demonstrated that the desire to continue education was among the most popular reasons for girls to opt for an abortion (25,30). Participants’ opinions also varied on whether abortion should be allowed in cases where the mother does not want to keep the baby. Only a small proportion of participants recognized access to safe abortion as a woman’s reproductive right, thus shedding light on most participants’ views on sexual and bodily autonomy. Failure to recognize abortion as a reproductive right undermines the widespread protection of abortion under the rights to life, as well as global recognition of access to safe, legal abortion as a fundamental right of women (31,32). This warrants a need for further education on the sexual and reproductive health rights of girls and women in Namibia.

Most participants showed strong support for abortion to be permitted in cases of rape and in conditions where the pregnancy poses a health risk to the woman. Similarly, studies in Nigeria found that participants were in support of legalizing abortion on medical grounds and in cases of rape or incest (21,33). These perceptions were in line with the current abortion laws in Namibia that stipulate that abortion may be permissible when the continuation of the pregnancy endangers the mother’s life, when the child is at risk of severe physical and mental defect, and when the pregnancy results from rape or incest (34). Participants’ beliefs, however, appeared to be divided on whether abortion should be permissible if the pregnant woman is mentally incapacitated or if the fetus has congenital anomalies. According to a report by the Guttmacher Institute, abortion is not permissible under any circumstances, in 14 African countries and is generally permitted only to save the life of the woman (35,36). While we did not conduct an analysis of the differences between these viewpoints, the perceptions of the participants regarding the circumstances for permissible abortion may be reflective of participants’ openness to conditional, scenario-based abortions.

We also observed divided opinions among the participants regarding the intersections between abortion and contraceptive use. The stance of the participants on whether legalizing abortion would lead to women using abortion as a substitute for contraceptives or would undermine pregnancy prevention methods was inconclusive. There is currently a dearth of research on trends in contraceptive use following abortion legalization in Africa and other countries. In a review of abortion and contraceptive use in Sub-Saharan Africa, Don Laura posits, based on the available evidence, that the contemporary use of abortion in Sub-Saharan Africa often substitutes for and sometimes surpasses modern contraceptive practices (36). The review further concluded that abortion has been and will likely continue to be used as a means of family planning within much of sub-Saharan Africa (36). It is, however, plausible that the increased use of abortion may be reflective of the unmet need for contraceptives, especially among adolescents. While Namibia has made significant strides in increasing the contraceptive prevalence rate (CPR) from 23% in 1992 to 61% in 2020 (37) teenage pregnancy rate remains a serious concern, with one in every five (19%) girls between the ages of 15 to 19 years becoming pregnant (37). Unintended pregnancies push adolescents to resort to clandestine, unsafe abortions, which increases the risk of health complications such as sepsis, severe anemia, disabilities, infertility and death (29,30).

Participants also had divided opinions on whether legalizing abortion would reduce the social stigma surrounding illegal abortion. Abortion stigma is a widely recognized contributor to secret unsafe abortions, often due to the strong social construction of the immorality of abortion (22,30). A systematic review by Munakampe et al (2018) found that the fear of shame following an abortion was far much higher than the stigma due to pregnancy, which could potentially explain why adolescents would not opt for safe abortions even in places where abortion is legal (30). These results could be suggestive that legalizing abortion and increasing access to safe abortion might not necessarily reduce unsafe abortion practices. Further research is needed to investigate the relationship between legalizing safe abortions and incidences of illegal, unsafe abortions.

### Strengths and limitations

Our findings should be considered in the context of the strengths and limitations of our study population and design. First, the study only captured the perceptions of women residing in //Gereres, Keetmanshoop, and therefore cannot be generalized for the entire Namibia. Second, the study only included women. Capturing the perspectives of men could be insightful in comparing the gender differences in perceptions regarding the legalization of abortion in Namibia. Third, we did not conduct statistical analysis to determine the correlation strength of the factors influencing the participants’ perceptions. In terms of strengths, our study is among the few studies conducted in Namibia to assess the perceptions on the acceptability of legalizing abortion. The study expands our understanding of the important factors to be considered in the legalization of abortion in Namibia.

## Conclusion

Our findings demonstrate that women were mostly not in support of legalizing abortion in Namibia. Religious and cultural beliefs appeared to be highly influential in the participant’s perceptions regarding abortion legalization. Participants’ concerns that abortion legalization would lead to promiscuity and its potential implication in increasing sexually transmitted diseases were found to be important factors. The study also highlighted that participants had varied opinions about the circumstances under which abortion should be permissible but demonstrated openness for inclusivity regarding conditions for permissible abortion. Moreover, the study shed light on fears and concerns about abortion being used as a substitute rather than complementing contraceptive use and other family planning methods. Further research is needed to explore the health policy implementation strategies that can prevent unintended pregnancies, avert the negative consequences of illegal, unsafe abortions, and achieve the Sustainable Development Goals for improved maternal and adolescent health.

## Data Availability

Data can be requested from authors and will be made available immediately on request.

## Acknowledgements

We wish to thank all the authorities that granted us the permission to conduct this study and the Bachelor of Nursing students at the University of Namibia, Southern Campus and Main Campus who gave up their time to be involved in this study. Thank you.

